# A Rapid Method to Evaluate Pre-Travel Programs for COVID-19: A Study in Hawaii

**DOI:** 10.1101/2021.03.06.21251482

**Authors:** Amy T. Hou, Genevieve C. Pang, Kristin M. Mills, Krizhna L. Bayudan, Dayna M. Moore, Luz P. Medina, Lorrin W. Pang

## Abstract

**Background:** Pre-travel testing programs are being implemented around the world to curb COVID-19 and its variants from incoming travelers. A common approach is a single pre-travel test, 72 hours before departure, such as in Hawaii; however this raises concerns for those who are incubating or those infected after pre-travel testing or during transit. We need a rapid method to assess the effectiveness of pre-travel testing programs, and we use Hawaii as our case study.

**Methods:** We invited travelers departing from Kahului main airport at the end of their visit to Maui (major tourist destination among the Hawaiian islands) and performed COVID-19 PCR testing. Eligible participants needed a negative pre-travel test and a Hawaiian stay ≤ 14 days. We designed for anonymous testing at the end of travel so that travel plans would be unaffected, and we aimed for ≥ 70% study participation.

**Results:** Among consecutive eligible travelers, 282 consented and 111 declined to participate, leading to a 72% (67-76%, 95% confidence interval) participation rate. Among 281 tested participants, two were positive with COVID-19, with an estimated positivity rate of 7 cases per 1,000 travelers. The top states of residence are California (58%) and Washington (21%). The mean length of stay was 7.7 ± 0.2 days. Regarding pre-travel testing, 87% had non-nasopharyngeal tests and 66% had self-administered tests.

**Conclusions:** This positivity rate leads to an estimated 17-30 infected travelers arriving daily to Maui in November-December 2020, and an estimated 52-70 infected travelers arriving daily to Hawaii during the same period. These counts surpass the Maui District Health Office’s projected ability to accommodate 10 infected visitors daily in Maui; therefore, an additional mitigation layer for travelers is recommended. This rapid field study can be replicated widely in airports to assess effectiveness of pre-travel programs and can be expanded to evaluate COVID-19 importation and its variants.

## Introduction

Communities face the challenge of regulating travel and tourism during COVID-19. Destinations with a high economic reliance on tourism have strategized to limit viral importation with health screens, quarantines, pre-travel testing, and post-arrival testing. At the start of the pandemic, Hawaii implemented universal 14-day quarantine for all travelers, resulting in a massive decline of visitors and the third lowest COVID-19 rate among the states.^1,2^ However, given its dependence on tourism, Hawaii established the Safe-Travels Program (STP) in October 2020 whereby travelers could submit a negative polymerase chain reaction (PCR) test by a trusted partner within 72 hours to bypass quarantine.^3^

A single pre-travel test raises some concerns: 1) travelers who were incubating SARS-CoV-2 and escaped detection at the time of testing, and 2) travelers who were infected by SARS-CoV-2 after pre-travel testing or during transit. Furthermore, it would allow the entry of travelers infected up to 3 days prior to pre-travel testing, given that the prepatent period for SARS-CoV-2 virus is estimated at 3 days, based on studies showing a median incubation of 5 days and peak viral shedding at 2 days prior to symptoms onset.^4–8^ Given this prepatency, the STP guidelines may still permit such travelers to enter and transmit the virus within the community.

In an effort to assess Hawaii’s STP program, a study was initially conducted and reported a COVID-19 positivity rate of 0.65 cases per 1,000 travelers;^9^ however, it had less than 10% participation rate and its methodology had concerns for self-deselection and distortion bias. Reviewers critiqued its validity and projected a revised positivity rate as high as 7-15 per 1,000 travelers.^10^ We adapted and improved our approach towards more robust sampling with randomized, consecutive solicitation followed by high participation rates (>65-70%),^11^ estimating an appropriate sample size and limiting detracting factors.

The effectiveness of a single pre-travel test is still uncertain. Some propose the addition of a post-arrival test with or without quarantine; while others propose a return to universal quarantine. Mathematical models can be used to predict the impact of various mitigation towards COVID-19; such as: risk reduction by 37-61% with departure-day testing, 97-100% with a 14-day quarantine after arrival, or 95-99% with a 7-day quarantine after arrival and post-arrival test on day 3-4.^12^ Worldwide, a rapid field method is needed to validate models and evaluate pre-travel testing programs, particularly with the emerging variants.

Prior to the study, focus groups were conducted to optimize participation. Eight groups of 15-20 mainland arrivals at Maui Kahului airport (OGG) helped to determine that the best time for the study was at the end of travel to avoid impact on travel plans, to test at airport departure gates (rather than clinics or hotels), and that positive results would not be reported to home states. It was important to them that accurate tests were used, and that results would be made known to them within days.

## Methods

The Maui District Health Office (Department of Health), Department of Transportation, and Maui County Medical Society partnered to conduct the study and enrolled participants from November 20-30, 2020 at five departure gates in Maui Kahului airport. Individuals were invited to participate if they had a primary residence outside of Hawaii, traveled from a location other than Hawaii, had a negative pre-travel PCR test 72 hours before departure, and stayed in Hawaii for up to 14 days. Participants were recruited in an active, sequential manner, and participation was limited to one person per household or travel group. Persons who had a primary residence in Hawaii or stayed greater than 14 days were excluded. Individuals who were interested but had inadequate time before boarding were also excluded. Study participants answered a brief survey with mandatory questions (state of residency and any locations visited 14 days prior to Hawaii) and several optional questions for contact tracing, including lodging, restaurants and any COVID-19 symptoms.

Each study participant underwent a nasopharyngeal (NP) PCR test, performed by a medical staff, and then was given a complimentary Hawaiian-designed face mask. On study day 5, investigators added data collection at the time of the NP swab regarding the pre-travel PCR testing: NP or non-NP (i.e. nasal, oropharyngeal and salivary) swabbing; and whether it was self-administered. The COVID-19 test was provided free to participants, with results available in 24-72 hours, were anonymous/confidential, were not to affect travel plans and not reported to home states or airlines. Each participant was given a random, unique study number. Results were posted on a website with a coded positive or negative result only decipherable by the participant, in order to assure confidentiality of each result and the aggregate positivity rate. Participants also had the option to receive results via text messaging.

Using the Clopper-Pearson (CP) Method^13^ and our goal of at least 70% participation, we estimated the target number of eligible individuals to solicit for the study and to detect positive COVID-19 cases (Table 1 and 2). For analysis, the positivity rates would be multiplied by the number of daily arriving visitors (obtained from Hawaii tourism data from the Department of Transportation) to estimate the number of infections entering on a daily basis.

**Table 1.** Point Estimates of Participation Rates Using a Total of 400 Solicited Individuals with Varying Number of Participants Who Enroll.

| # of Participants Who Enroll into Study | Point Estimate of Participation Rate | 95% CI of Participation Rate |
| --- | --- | --- |
| 360 | 90% | (87% - 93%) |
| 320 | 80% | (76% - 85%) |
| 280 | 70% | (65% - 74%) |
| 240 | 60% | (55% - 65%) |
| 200 | 50% | (45% - 55%) |

**Table 2.** Point Estimates for a Valid Representative Population Using Study Size of N = 300.

| # Positive PCR Tests<br>in Sample Study | Point Estimate of Overall Covid Cases<br>(per 1000) | 95% CI<br>(per 1000) |
| --- | --- | --- |
| 0 | 0 | (0 - 17) |
| 1 | 3 | (0.1 - 18) |
| 2 | 6 | (0.8 - 24) |
| 3 | 9 | (2 - 30) |
| 5 | 16 | (5 - 38) |
| 7 | 23 | (9 - 38) |
| 10 | 33 | (16 - 60) |

This study has been reviewed by the Institutional Review Board of the Hawaii State Department of Health, which approved that the study met the criteria for public health surveillance based on 45 CFR 46.102 (l)(2) of the Department of Health and Human Services.

## Results

Among 577 individuals screened during the study period, 184 were excluded, 282 agreed to participate and 111 declined, as described by Figure 1. Of those eligible, the participation rate was 72% (67-76%, 95% confidence interval [CI]).

**Figure 1.**
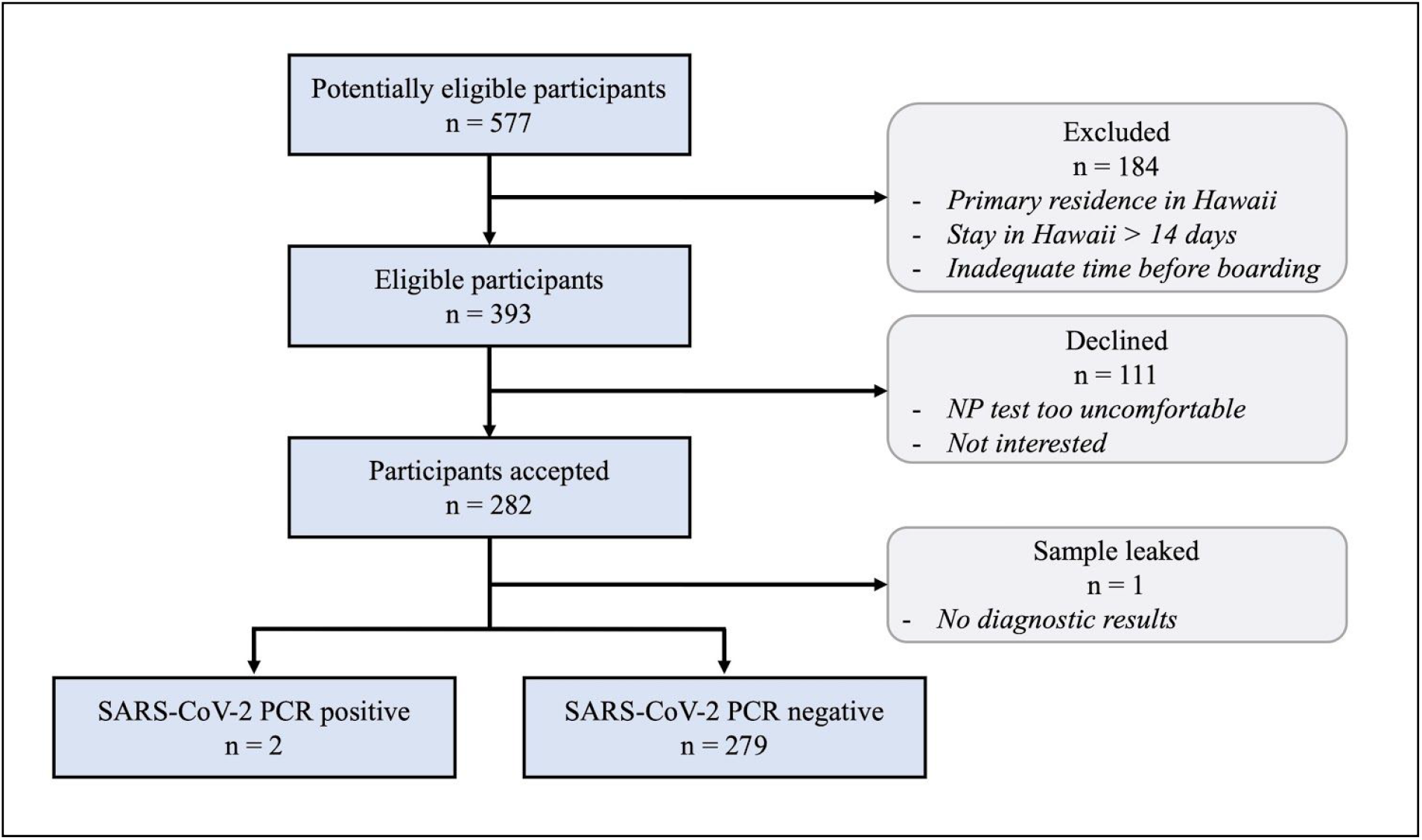
Flow Diagram of Study Participants.

Table 3 describes the demographic characteristics of the study participants. The top primary states of residence for study participants were California (58%), Washington (21%) and Colorado (8%), reflecting the departure gates assigned to this study. The mean length of stay was 7.7 ± 0.2 days. The distribution of the lengths of stay is displayed in Figure 2 clustering between 5-10 days, which was preferred to increase the chances of capturing travelers that had incubating or prepatent periods.

**Table 3.** Demographic Characteristics of Study Participants.

| **Mandatory Questions** | **Study Group** |
| --- | --- |
| State of residence (N=281) — no. (%) |  |
| California | 164 (58) |
| Washington | 58 (21) |
| Oregon | 22 (8) |
| Colorado | 4 (1) |
| Nevada | 4 (1) |
| Arizona | 2 (1) |
| Arkansas | 2 (1) |
| Wisconsin | 2 (1) |
| Other | 23 (8) |
| Number of days in Hawaii (N = 272) |  |
| Range (days) | 0 - 14 |
| Mean (days) | 7.7 ± 0.2 |
| **Optional Questions** |  |
| Answered any optional question — no. (%) | 270 (96) |
| Provided phone number for results — no. (%) | 173 (62) |
| Negative pre-travel test (N = 218) — no. (%) |  |
| 3 days prior to departure | 165 (76) |
| 2 days prior to departure | 47 (21) |
| 1 day prior to departure | 6 (3) |
| Any travel prior to arriving in Hawaii? (N =270) — no. (%) |  |
| Yes | 14 (5) |
| No | 256 (95) |
| Those who traveled to Oahu before Maui (N = 4) |  |
| Mean stay in Oahu (days) | 3 |
| Lodging in Maui (N = 233) — no. (%) |  |
| Hotel | 149 (64) |
| Condo/timeshare | 61 (26) |
| House | 23 (10) |
| Did excursion tour, no. | 50 |

**Figure 2.**
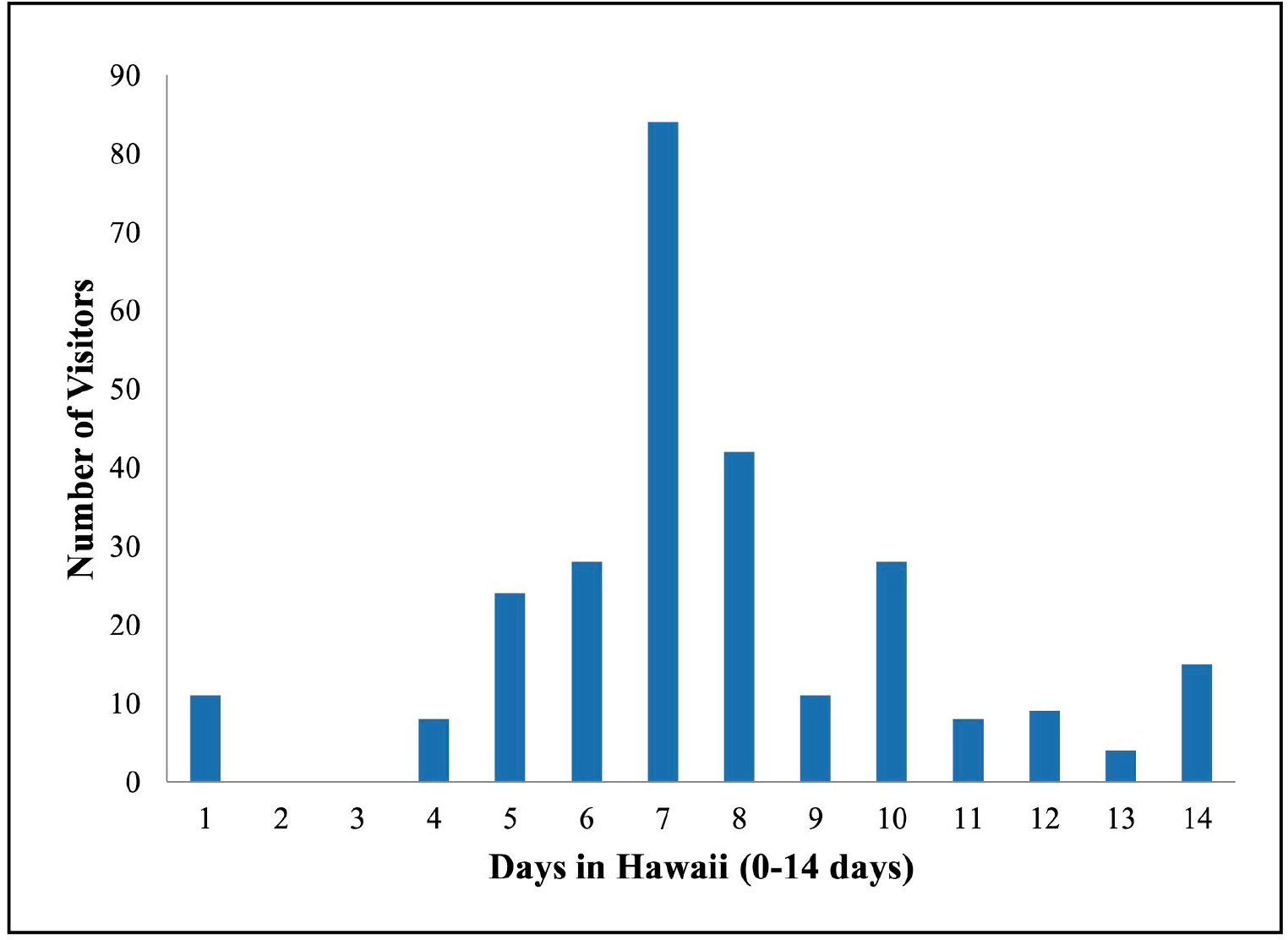
Distribution of Visitors by Lengths of Stay in Hawaii (N = 272)

Among participants who answered questions regarding accommodations, the most frequently reported were hotels (64%) and condominiums/timeshares (26%), located in the popular areas of West and South Maui. Only 14 participants traveled elsewhere within the two weeks before arrival to Hawaii, and only four reported going to Oahu before Maui for a mean stay of three days. One participant reported gastrointestinal symptoms that could be compatible with COVID-19 but tested negative for COVID-19.Among participants, 96% answered optional questions, and 62% provided phone numbers for results, therefore the additional questions did not appear to deter participation.

Two subjects tested positive and 279 negative for COVID-19, resulting in a positivity rate of 7 cases (1-24, 95% CI) per 1,000 visitors. One positive asymptomatic case traveled from California, had a negative pre-travel test 3 days prior to arriving in Maui, and stayed at a hotel for 7 days. The second positive asymptomatic case traveled from Wisconsin with a 1-day stay in California, had a negative pre-travel test 3 days prior to arriving to Maui, and stayed on Maui for only one day (unclear if this case was departing to another Hawaiian island). One enrolled participant’s sample leaked before diagnostic testing, and the test was discarded. Table 4 illustrates data collected on the pre-travel testing from participants.

**Table 4.** Pre-Travel COVID-19 Testing Characteristics.

|  | Nasopharyngeal | Not Nasopharyngeal |
| --- | --- | --- |
| Self-Administered | 3 | 132 |
| Not Self-Administered | 22 | 48 |

## Discussion

Our rapid assessment of Hawaii’s STP and the COVID-19 positivity rate among travelers incorporated representative sampling with active, randomized, sequential recruitment, combined with a high 72% participation rate. Our data suggested a positivity rate of 7 cases per 1,000 travelers. Applying this positivity rate to the available tourism data derives an estimate of 17-30 infected travelers arriving daily to Maui in November-December 2020, and an estimate of 52-70 infected travelers arriving daily to Hawaii state in the same period.^3^ These counts surpass the Maui District Health Office’s projected ability to care for 10 infected visitors per day in Maui, based on its low community incidence and hospital capacity. Since December 2020, the rising number of visitors and increased transmission on the mainland is likely associated with an even higher number of cases currently entering Hawaii.

The other Hawaii travel study (Miller) described a significantly lower positivity rate of 0.65 cases per 1,000 travelers; it was conducted from mid-October to November 2020 on several islands: Maui, Oahu, Kauai and Big Island.^9^ However, its validity has been challenged due to several factors: 1) it notified 10% of arriving passengers via online email, inviting voluntary participation in post-arrival testing 2) it allowed for the less-accurate rapid antigen testing; and 3) it reported results to health officials who imposed a 10-day isolation period on those testing positive and a 14-day quarantine on their close contacts, plus a no-fly notification to airlines that was devastating to travel plans. The methodology was associated with a low participation rate (<10% of those invited) challenged by self-deselection and a distortion bias that cannot be compensated, even by enrolling high numbers of study participants. Our study design aimed to mitigate these observed disincentives and improve participation.

The two positive cases identified in our study were most likely infected in their primary residential states of California and Wisconsin; since, during the study period, COVID-19 case rates were 14-fold higher in California and 8-fold higher in Wisconsin, than for Hawaii state,^2^ and even higher compared to Maui, a lower transmission district at the time.^14^ It is plausible for the first positive case to have been exposed in Maui rather than California given the median incubation period of 5 days and this case’s stay of 7 days; however, rates were far higher in California.

It is important for Hawaii to acknowledge the estimated introduction of 7 COVID-19 cases per 1,000 incoming mainland travelers. Given the potential COVID-19 spread to the local community and our limited hospital capacity, we recommend another layer of mitigation to the STP for incoming travelers, such as scientifically-based post-arrival testing with shortened quarantine.^12^ The Kauai COVID-19 Discussion Group has suggested adding a second test after 6 days of quarantine to attempt decreasing travelers’ positivity rate to <5 per 10,000.

For the pre-travel testing required by Hawaii, the majority of our participants (87%) did not undergo NP testing and many (66%) were self-administered, which may introduce the potential insensitivity of pre-travel COVID-19 testing.^15,16^ It is plausible that travelers opted for more convenient tests (i.e. self-administered) or more comfortable tests (i.e. non-NP). We did not pair the pre-travel test to each participant, therefore the types of pre-travel testing for the two positive cases were unknown. In future studies, collecting these details may help to determine the sensitivity among the many pre-travel tests.

Our study was limited in size: future studies could be performed with larger numbers to further tighten the confidence interval around point estimates, which for our purposes focused on participation rates to be about 70%. Our sample included mainly travelers from California and Washington, due to the departure gates where our study was stationed; this may potentially add some selection bias. We sought minimal information from participants in order to achieve high participation. We selected one member from each travel group to maximize sampling from different travel groups, but inclusion of other members of the two positive cases may help to investigate additional cases. There is a discrepancy in the total data collected for the lengths of Hawaii stays (mandatory question) due to incomplete data entries from the study day 1. We may have missed infections in participants who were infected but produced a negative result by the time of our study’s testing (particularly for those who stayed 10-14 days in Hawaii). However, the positivity rates estimated for travelers staying 0-14 days may be extrapolated to the entire traveler group, regardless of length of stay. Finally, we did not perform genomic analysis, which could establish molecular-level relatedness, and should be implemented in the future for detecting and measuring the rate of incoming COVID-19 variants.

This simple, rapid field study has implications for all pre-travel testing programs that aim to estimate the importation of COVID-19. For destinations with relatively low prevalence of a target, such as a specific variant, our approach can assess the positivity rate of the target in travelers. To estimate the introduction of COVID-19 variants, genomic testing will be needed, ideally multiple variant genomic testing on a single sample. In future studies, Hawaii can conduct this study at the main airports in Maui and Oahu, and add genomic testing, to track the positivity rate of COVID-19 and its variants into Hawaii. Recently, the CDC has called for a single pre-travel test in all travelers entering the United States;^17^ thus, this study can be readily utilized to assess positivity rates of travelers at a broader scale in our country. It would be useful to repeat this simple study periodically (such as every two to three months) at airports worldwide, to evaluate the efficacy of travel control programs in the face of rising COVID-19 infections and variants. This tool can assist communities in evaluating whether the travelers’ positivity rates exceed their health-system’s capacities to care for additional incoming cases, and whether to consider adding additional layers of mitigation for travelers.

## Data Availability

All authors confirm that they had full access to all the data in the study.

## Acknowledgments

Anna Metcalfe, Ryan Tomashiro, Priscilla Kalani Holokai, Kathleen Shida-Pang, Olivia Jenkins, John Cheetham, Robert Santry, Norman Estin, Belia Paul, Cara Flores, Cole Whitney, Shannon McCaffrey, Lin H. Chen

